# The urgency of resuming disrupted dog rabies vaccination campaigns: a modeling and cost-effectiveness analysis

**DOI:** 10.1101/2021.04.24.21256032

**Authors:** Amber Kunkel, Seonghye Jeon, Haim C Joseph, Pierre Dilius, Kelly Crowdis, Martin I. Meltzer, Ryan Wallace

**Author notes:** **Disclaimer:** The findings and conclusions in this article are those of the authors and do not necessarily represent the official position of the U.S. Centers for Disease Control and Prevention (CDC).

## Abstract

**OBJECTIVE:** Dog vaccination is a cost-effective approach to preventing human rabies deaths. In Haiti, the 2019 dog vaccination campaign did not include the capital city, and the 2020 campaign was cancelled because of COVID-19 lockdown restrictions and redirection of funds. We estimated the number of human lives that could be saved by resuming dog vaccination in 2021 compared to 2022 and compared the cost-effectiveness of these two scenarios.

**METHODS:** We modified a previously published rabies transmission and economic model to estimate trends in dog and human rabies cases in Haiti from 2005-2025. We compared model outputs to surveillance data on human rabies deaths from 2005-2020 and animal rabies cases from 2018-2020. We then estimated the human health and cost implications of restarting dog vaccination programs in either 2021 or 2022.

**FINDINGS:** Model predictions and animal surveillance data from Haiti both suggest a 5-to 8-fold increase in animal rabies cases has occurred in the capital city between Fall 2019 and Fall 2020. We estimate that restarting dog vaccination in Haiti in 2021 compared to 2022 could save 285 human lives and prevent 6,541 human rabies exposures over a five-year period and may decrease program costs due to reduced need for human post-exposure prophylaxis.

**CONCLUSIONS:** A one-year delay in resuming dog vaccination in Haiti, from 2021 to 2022, could cost hundreds of lives over the next 5 years. Interruptions in dog vaccination campaigns before elimination is achieved can lead to significant human rabies epidemics if not promptly resumed.

## BACKGROUND

Rabies is a neglected disease with a nearly 100% fatality rate. Dog rabies vaccination is a cost-effective strategy to prevent human rabies exposures and deaths ^1-3^. However, vaccination campaigns need to be repeated annually with at least 70% coverage among the susceptible dog population until elimination is achieved ^4-7^. Vaccination of humans with exposure to suspected rabid animals (post-exposure prophylaxis, or PEP) is also effective, but it is more expensive than dog vaccination and requires robust surveillance and integration between animal and human healthcare systems ^8,9^.

Haiti is one of only four countries in the Western Hemisphere that reports dog-mediated human rabies deaths. Expansions in Haiti’s dog vaccination programs, community bite surveillance systems, and PEP access have increased rabies case detection in dogs 18-fold ^10^ and decreased human rabies deaths by over 50% ^11^ since 2013. In 2019, Haiti reported just 2 human rabies deaths. However, this progress is now threatened. In 2019 nearly 500,000 dogs were vaccinated against rabies, but funding limitations halted the campaign before reaching Ouest Department, which is home to Haiti’s capital Port-au-Prince and nearly 4 million inhabitants. Haiti’s 2020 dog vaccination campaign was cancelled and funds were diverted to support the COVID-19 response; funds for 2021 have not yet been identified. Similar COVID-19 related postponements to dog vaccination campaigns have also been reported in other countries ^12^.

The cessation of dog rabies vaccination programs raises concerns that Haiti could soon be facing a major dog-mediated rabies epidemic. We compared animal rabies surveillance data with model-predicted trends in dog rabies cases from 2018-2020 in Haiti. We then compared the human lives potentially saved and the cost-effectiveness of restarting dog vaccination programs in either 2021 or 2022. These results show the impact that even short-term (2018-2021) interruptions to dog vaccination campaigns could have on human rabies deaths and progress towards elimination goals.

## METHODS

We modified the previously published RabiesEcon model to estimate dog and human rabies deaths in Haiti from 2021-2025 under several vaccination program scenarios. We obtained input values and ranges from the literature and unpublished program data from Haiti. We adjusted the most uncertain parameter values within these ranges so that model estimates would match approximate trends in human rabies deaths reported for 2005-2020 (Supplementary Appendix Tables S1 and S2). We assumed that 10% of human rabies deaths in Haiti are detected through current surveillance systems, a value that is consistent with estimates for other lower and middle income countries; even in upper income countries, approximately 50% of human rabies cases may be misdiagnosed ^13-15^.

To further validate the model, we compared trends in model-estimated dog rabies cases from July 2018-November 2020 to animal rabies surveillance data. We then used the model to estimate future human rabies deaths for the period 2021-2025, assuming dog vaccination programs are resumed in either 2021, 2022, or never resumed. We estimated the cost-effectiveness of these vaccination scenarios compared to a baseline scenario of no dog vaccination and no PEP for 2021-2025.

### Model

RabiesEcon is a spreadsheet-based tool that models the spread of rabies among dogs using a susceptible, exposed (via bites from infected dogs), infected/ infectious, and removed/immune (SEIR) framework. Humans become exposed through the bite of a rabid dog. Exposed humans who do not receive PEP are at risk of developing rabies, with a mortality rate of 100%.

The original model was modified to allow for year-to-year changes in dog vaccination coverage. Vaccination campaigns were assumed to occur in the middle of each year and last for 10 weeks, with coverages defined by historical data and intervention scenarios.

We ran separate versions of the model for Ouest Department and the rest of Haiti. We added the results to create country-wide estimates and did not address the potential movement of dogs between regions. Ouest Department is the home of Port-au-Prince, the capital city, and more than one third of Haiti’s inhabitants. However, funding limitations prevented its inclusion in Haiti’s 2019 dog vaccination campaign; the last vaccination campaign in Ouest occurred in early 2018.

We used a time horizon of five years (2021-2025) for all analyses. We computed total program costs for three intervention scenarios with PEP coverage maintained at current levels: resuming dog vaccination in 2021, resuming dog vaccination in 2022, and never resuming dog vaccination. We compared these to a “no intervention” scenario with no dog vaccination and no PEP. The five-year average cost-effectiveness ratios per human death averted were calculated for each scenario by dividing total program costs by the reductions in human rabies deaths compared to the no intervention scenario.

### Data and input values

We used the same values of most rabies natural history parameters as previously published versions of the model (Supplementary Appendix Table S1). However, we assumed a decay of immunity after dog vaccination of 0.0096 per week, equivalent to an average duration of immunity of 2 years following primary vaccination ^3,16,17^. To avoid artificially inflating vaccination coverage, we assumed vaccination campaigns stop once the total proportion of immunized dogs (either newly vaccinated or previously vaccinated and still immune) reaches the desired vaccination coverage. Essentially, we assumed vaccination campaigns reach all previously vaccinated dogs first before reaching susceptible dogs with no history of vaccination. Vaccination coverage for 2005-2020 was based on data from past vaccination campaigns (Supplementary Appendix Table S3). Once vaccination campaigns are restarted, we assume that they will recur annually with 70% coverage (a target met or exceeded in the 2018 and 2019 campaigns).

Haiti-specific input values were based on both published and unpublished data (Supplementary Appendix Table S1). We removed very low-density areas from population density estimates, as such regions may be unable to sustain rabies transmission cycles in the absence of continued importation (see Supplementary Appendix for details). We assumed 85% PEP coverage in Ouest Department and 80% coverage outside of Ouest ^11^. We also assumed that 5 total people are treated with PEP for each truly exposed case (Supplementary Appendix Table S1).

The total program costs for each scenario were calculated by adding the costs of community bite surveillance (such as lab tests and bite investigations), dog vaccination, and human PEP. We assumed average costs of $10.14 per suspect cases investigated ^18^, $72.91 for PEP costs ^18^, and $2.92 per dogs vaccinated (data not yet published) (Supplementary Appendix Table S1).

We adjusted input parameters so that model estimates would approximately match trends in surveillance-detected human rabies deaths in Haiti from 2005-2020 (Supplementary Appendix Table S1-S3), assuming that 10% of human rabies cases in Haiti are detected through surveillance ^13-15^. Human rabies surveillance in Haiti is conducted by the Ministry of Public Health and Population and is based primarily on hospital-based reporting of clinically suspected human rabies cases.

To further validate model outputs and assess the current rabies situation in Haiti, we compared model outputs of dog rabies cases from July 1, 2018-December 31, 2020 with animal surveillance data for the same period. We obtained surveillance data on animals investigated for rabies in Haiti under the Ministry of Agriculture, Natural Resources and Rural Development’s Haiti Animal Rabies Surveillance Program (HARSP) from July 1, 2018-November 19, 2020. Dogs and other animals that have been found dead or are suspected of rabies due to illness or behavior are reported to HARSP by veterinarians, healthcare workers, or members of the public ^10^. A standardized electronic case investigation form is used to document investigations, and provides near real-time information on epidemiologic changes ^19^.

### Sensitivity analyses

In sensitivity analyses we assumed 1-10 total people receive PEP per true rabies exposure. Furthermore, the RabiesEcon model has been shown to be most sensitive to values of the dog birth rate, dog life expectancy, and basic reproductive number, R0 ^20^. Therefore, we considered two additional scenarios: a high turnover scenario using higher-than-baseline values for dog birth and death rates (which define dog turnover rate) and lower R0 (number dogs infected per infectious dog); and, a low turnover scenario using lower values defining dog turnover but a higher R0 (Supplementary Appendix Table S2). These parameter combinations were selected to produce similar trends in human rabies deaths from 2005-2020 as the baseline scenario (Supplementary Appendix Fig S1-S3).

## RESULTS

### Dog rabies cases, 2018-2020

We compared model estimates of the number of dog rabies cases in 6-month intervals (Figure 1, left hand side) with surveillance data on the number of confirmed rabid animals (Figure 1, right hand side). The comparisons are shown separately for Ouest Department and outside Ouest Department.

**Figure 1:**
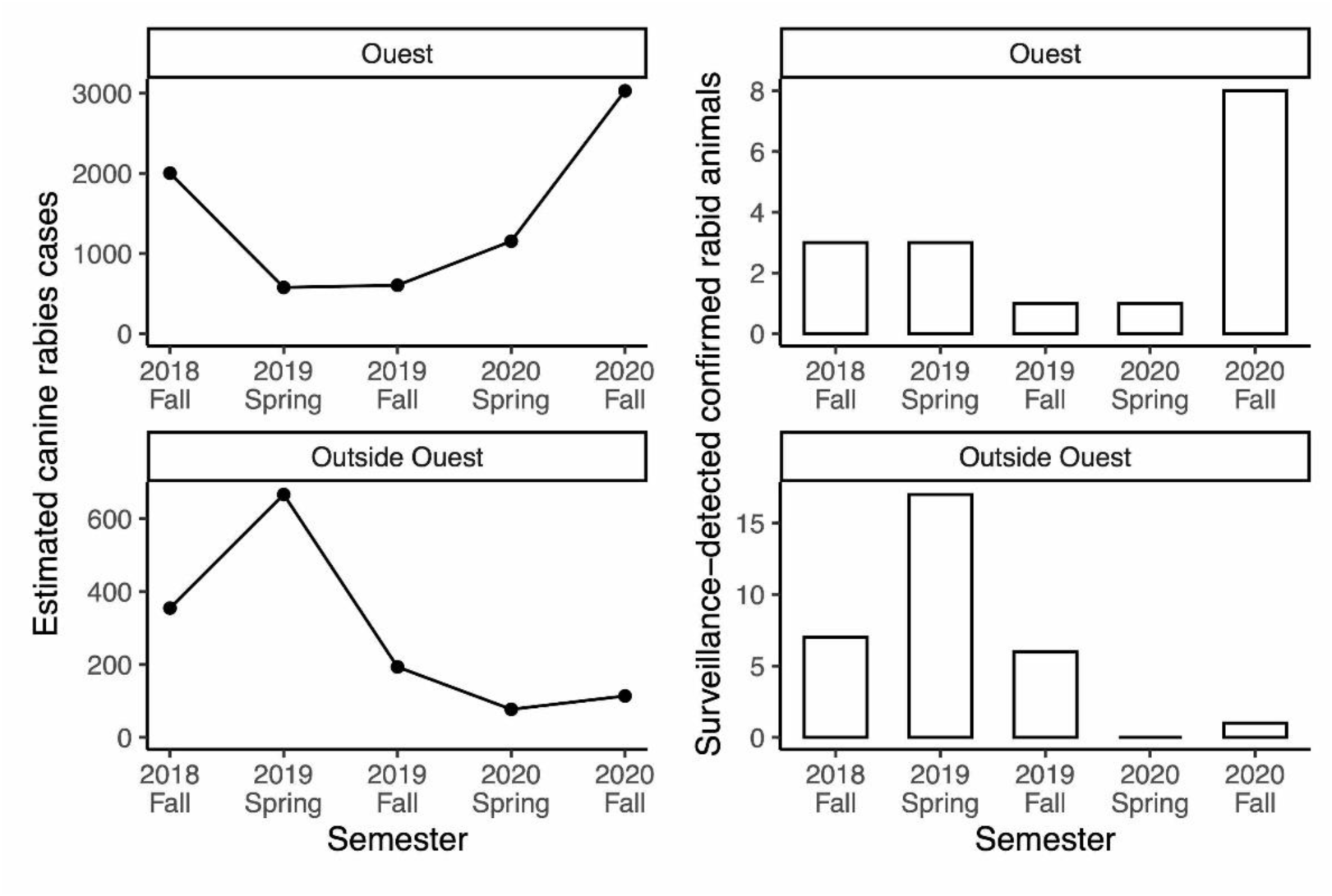
Dog rabies cases in Haiti since July 2018, as total number of cases estimated by the model*† (left) and percentage of all surveillance investigations resulting in confirmed positive cases (right). * Results are presented separately for Ouest Department (top), where the last vaccination campaign took place in 2018, and outside Ouest Department (bottom), where the last vaccination campaign took place in 2019. † These plots show estimated total dog rabies cases. Supplementary Appendix Table S4 compares model estimates of dog rabies cases and with confirmed rabid animals detected through surveillance data. Note that typically only animals with reported human bites are investigated.

In Ouest Department, where the last vaccination campaign occurred in 2018, dog rabies cases declined and remained low through 2019. However, there was a 5-8 fold increase in both model estimated and surveillance detected cases between Fall 2019 and Fall 2020 following the cessation of annual vaccination campaigns (Figure 1 and Supplementary Appendix Table S4). The estimated number of rabid dogs was 604 in fall of 2019 and 3,030 in fall of 2020. Outside of Ouest Department, a peak in cases occurred before the 2019 vaccination campaign. Model-estimated dog rabies cases subsequently declined to below 150 per six-month period through Fall 2020. The number of confirmed rabid animals per 6-month period ranged from 1-8 for Ouest Department and 0-17 for the rest of Haiti, suggesting case detection rates as low as 0.1% in Ouest Department (Supplementary Appendix Table S4).

### Predicted impact of delayed dog rabies vaccination from 2021 to 2022

We estimated that delaying dog vaccination from 2021 to 2022 would result in a human rabies epidemic with 285 excess deaths through 2025, of which 28 would be detected by the healthcare system (Figure 2). The corresponding number of dog rabies cases is shown in Figure S4.

**Figure 2:**
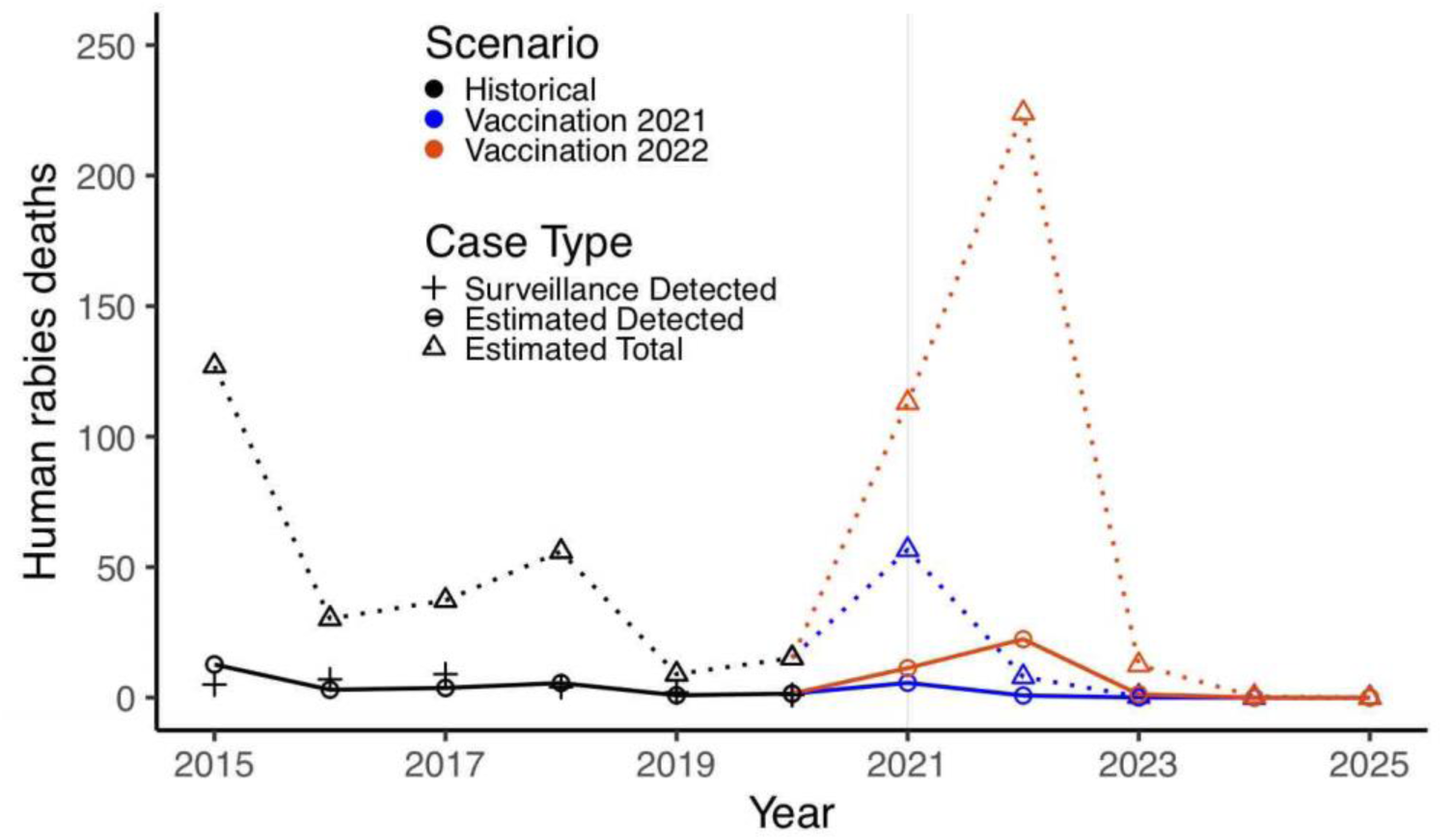
Historical and projected human rabies cases in Haiti, 2015-2025, and potential impact of delaying restarting dog rabies vaccination until either 2021 or 2022. * Black +’s indicate human rabies deaths detected by surveillance from 2015-2020. Circles (connected by solid lines) represent model estimates of detected human rabies deaths. Triangles (connected by dotted lines) represent model estimates of total human rabies deaths. We assume 10% of human rabies deaths are detected, which is consistent with estimates from other lower and middle-income countries ^13^. Colors compare historical data and estimates for 2015-2020 (black) with model predictions if vaccination campaigns resume in 2021 (blue) or 2022 (orange).

Restarting dog rabies vaccination in 2021 would have a five-year dog rabies vaccination cost of $7,530,776 (average annual cost $1,506,155) (Table 1), higher than the $5,757,508 if restarting vaccination were delayed until 2022. However, resuming dog vaccination in 2021 compared to 2022 would result in 63,050 fewer dog rabies deaths, 6,541 fewer human rabies exposures and 285 fewer human rabies deaths (of which 28 would be reported). It would also reduce the number of people needing PEP by 27,714 and lower PEP costs by $2,020,705 (Table 1). These effects would be even greater if dog vaccination is not resumed through 2025. Overall, restarting dog vaccination in Haiti in 2021 could lead to substantial cost savings of $284,794 compared to delaying until 2022, and $929,476 compared to no dog vaccination through 2025. (Table 1). With reference to a no intervention (no dog vaccination or PEP) scenario, resuming dog vaccination in 2021 would have an average cost per human death averted of $1,355, which is more cost-effective than either resuming dog vaccination in 2022 ($1,475 per human death averted) or performing no dog vaccination campaigns through 2025 ($1,950 per human death averted) (Table 1).

**Table 1:**
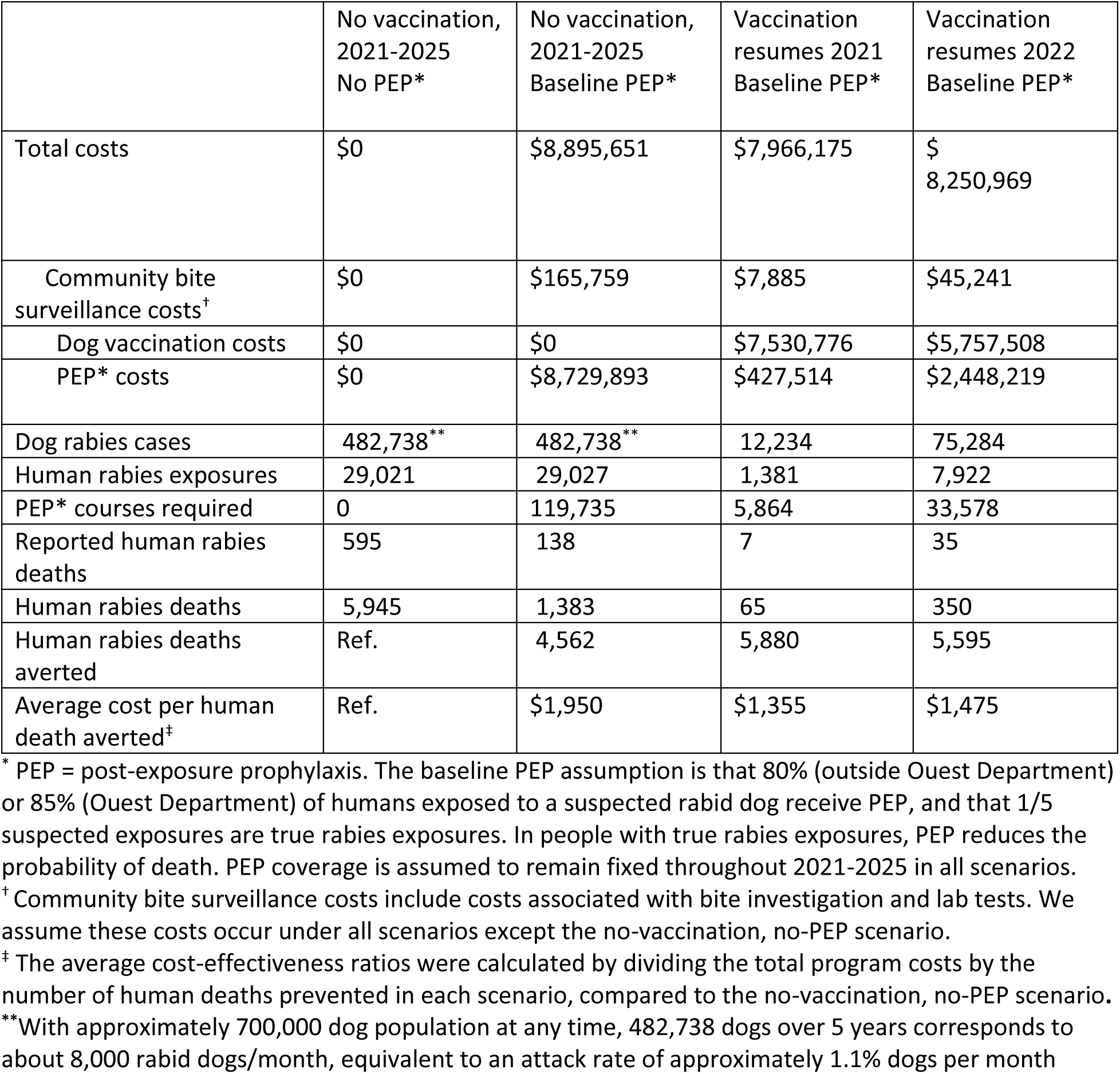
Five-year cost and health outcomes associated with combinations of dog rabies vaccination programs and human PEP*: 2021-2025.

### Sensitivity analyses

First, we compared different assumptions about PEP efficiency (Table 2). Resuming dog vaccination in 2021 is the most cost-effective scenario if at least 3 people receive PEP for every true rabies exposure. In a highly efficient healthcare system where fewer than 3 people get PEP per true exposure, providing PEP only would be more cost-effective. Under all scenarios, substantially more PEP is needed when vaccination is delayed past 2021 (5,500-55,000 additional courses over five years when delayed to 2022; 23,000-230,000 when delayed past 2025). The average cost-effectiveness ratio of restarting dog vaccination in 2021 and providing PEP (compared to no vaccination and no PEP) is similar across all assumptions about PEP use efficiency ($1,296-$1,429).

**Table 2:** Sensitivity analysis: Five-year health and cost effectiveness outcomes for different assumptions of PEP use efficiency.

| Outcome | Policy | Number of suspect rabies exposures per true rabies exposure** |  |  |  |
| --- | --- | --- | --- | --- | --- |
|  |  | 1 | 2 | 5 (Baseline) | 10 |
| Human deaths<sup>†</sup> | No vaccination<br>No PEP* | 5,945 | 5,945 | 5,945 | 5,945 |
|  | No vaccination<br>Baseline PEP | 1,383 | 1,383 | 1,383 | 1,383 |
|  | Vaccination 2021<br>Baseline PEP | 65 | 65 | 65 | 65 |
|  | Vaccination 2022<br>Baseline PEP | 350 | 350 | 350 | 350 |
| PEP*<br>courses<br>required | No vaccination<br>No PEP | 0 | 0 | 0 | 0 |
|  | No vaccination<br>Baseline PEP | 23,947 | 47,894 | 119,735 | 239,469 |
|  | Vaccination 2021<br>Baseline PEP | 1,173 | 2,345 | 5,864 | 11,727 |
|  | Vaccination 2022<br>Baseline PEP | 6,716 | 13,431 | 33,578 | 67,157 |
| Total<br>costs | No vaccination<br>No PEP | \$0 | \$0 | \$0 | \$0 |
| | No vaccination<br>Baseline PEP | \$1,779,130 | \$3,558,260 | \$8,895,651 | \$17,791,302 |
| | Vaccination 2021<br>Baseline PEP | \$7,617,856 | \$7,704,936 | \$7,966,175 | \$8,401,574 |
| | Vaccination 2022<br>Baseline PEP | \$6,256,200 | \$6,754,892 | \$8,250,969 | \$10,744,429 |
| Average<br>cost per<br>human<br>death<br>averted | No vaccination<br>No PEP | Ref | Ref | Ref | Ref |
| | No vaccination<br>Baseline PEP | \$390 | \$780 | \$1,950 | \$3,900 |
| | Vaccination 2021<br>Baseline PEP | \$1,296 | \$1,310 | \$1,355 | \$1,429 |
| | Vaccination 2022<br>Baseline PEP | \$1,118 | \$1,207 | \$1,475 | \$1,920 |

<sup>†</sup> Total human rabies deaths; we assume only 10% of these would be reported
\* PEP = post-exposure prophylaxis. The baseline PEP assumption is that 80% (outside Ouest Department) or 85% (Ouest Department) of humans exposed to a suspected rabid dog receive PEP, and that 1/5 suspected exposures are true rabies exposures. In people with true rabies exposures, PEP reduces the probability of death. PEP coverage is assumed to remain fixed throughout 2021-2025 in all scenarios.
\*\* The number of suspect rabies exposures per true rabies exposure is a measure of PEP use efficiency. A value of 1 indicates that only individuals with exposure to animals that are truly rabid receive PEP. Increasing levels imply decreasing PEP use efficiency.

We then compared the three parameter combinations described in the Methods, under the baseline assumption of PEP efficiency. In each scenario (Table S2), a large rabies epidemic is expected to begin in 2021. The number of excess human deaths associated with delaying the next dog vaccination campaign from 2021 to 2022 was predicted to be 107 under the “high turnover” scenario and 217 under the “low turnover” scenario, compared to 285 under the baseline scenario (Table 3 and Figure 3). The average five-year cost-effectiveness ratio per human death averted is similar when vaccination is resumed in 2021 compared to 2022 under both the “high turnover ($1,651 in 2021 vs. $1,510 in 2022) and “low turnover” ($1,323 in 2021 vs. $1,377 in 2022) scenarios. (Table 2).

**Table 3:**
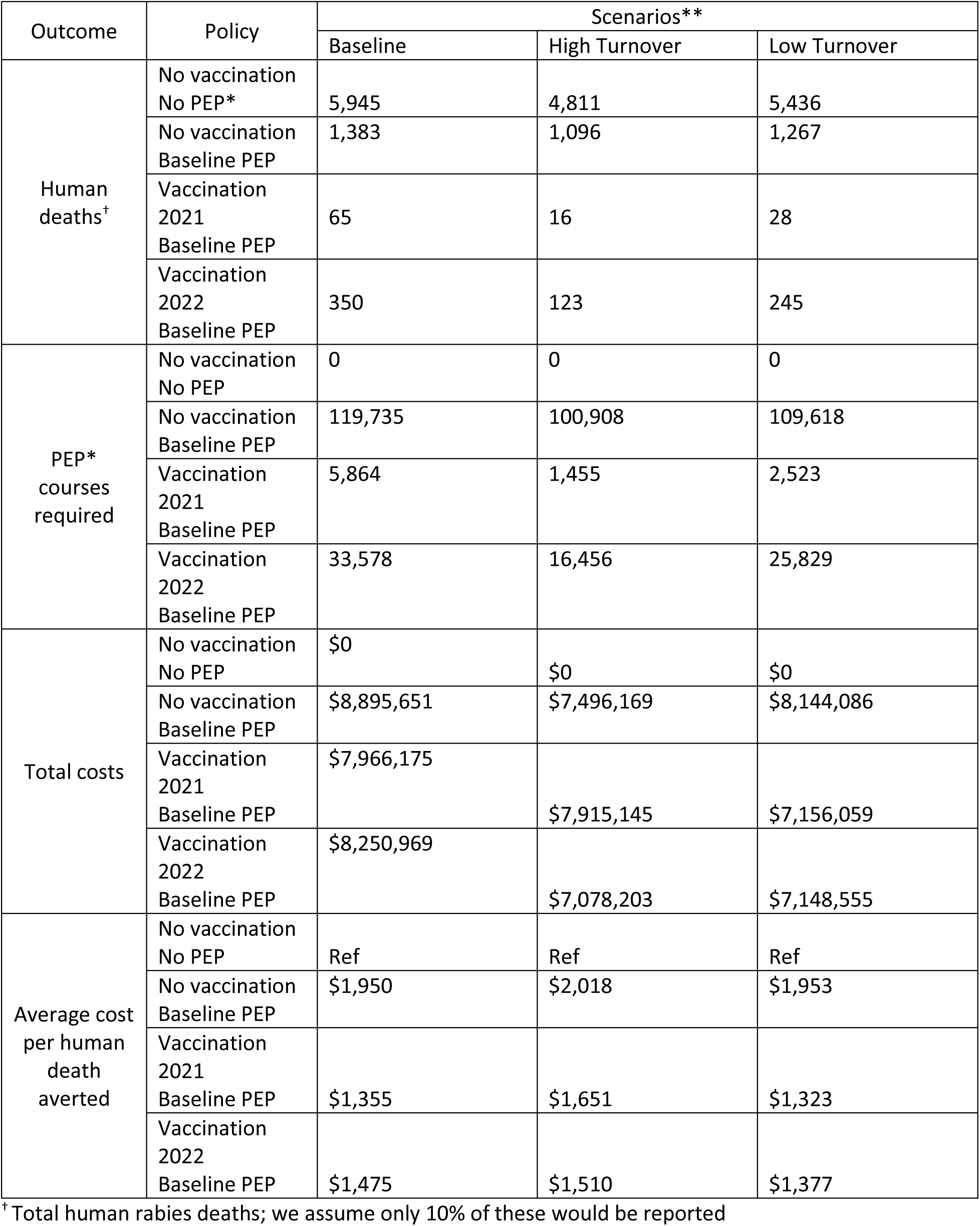
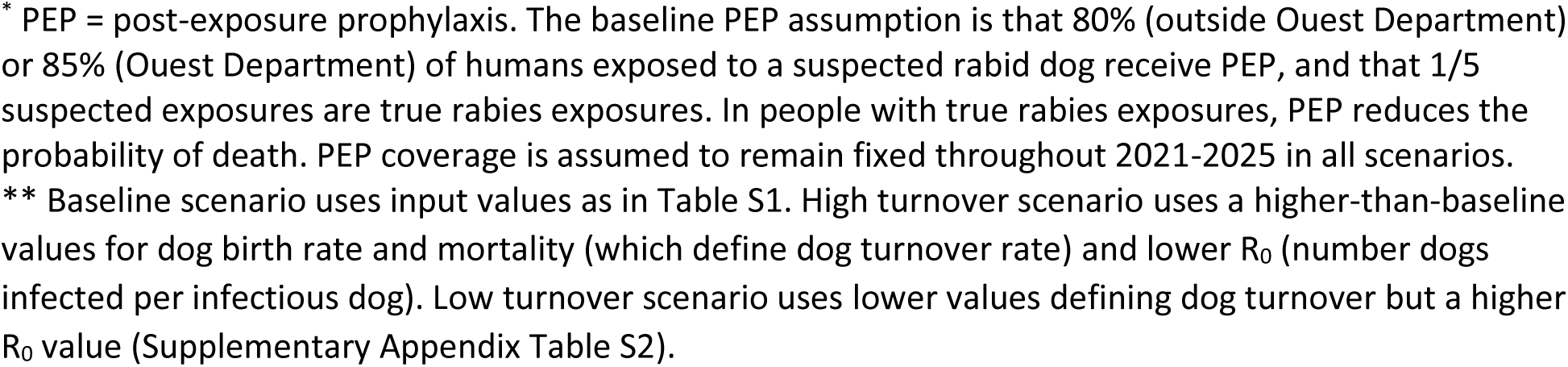
Sensitivity analysis: Five-year health and cost effectiveness outcomes for different scenarios of dog turnover and dog-to-dog rabies infectiousness.

| Outcome | Policy | Scenarios** |  |  |
| --- | --- | --- | --- | --- |
|  |  | Baseline | High Turnover | Low Turnover |
| Human deaths<sup>†</sup> | No vaccination<br>No PEP* | 5,945 | 4,811 | 5,436 |
|  | No vaccination<br>Baseline PEP | 1,383 | 1,096 | 1,267 |
|  | Vaccination<br>2021<br>Baseline PEP | 65 | 16 | 28 |
|  | Vaccination<br>2022<br>Baseline PEP | 350 | 123 | 245 |
| PEP*<br>courses<br>required | No vaccination<br>No PEP | 0 | 0 | 0 |
|  | No vaccination<br>Baseline PEP | 119,735 | 100,908 | 109,618 |
|  | Vaccination<br>2021<br>Baseline PEP | 5,864 | 1,455 | 2,523 |
|  | Vaccination<br>2022<br>Baseline PEP | 33,578 | 16,456 | 25,829 |
| Total costs | No vaccination<br>No PEP | \$0 | \$0 | \$0 |
| | No vaccination<br>Baseline PEP | \$8,895,651 | \$7,496,169 | \$8,144,086 |
| | Vaccination<br>2021<br>Baseline PEP | \$7,966,175 | \$7,915,145 | \$7,156,059 |
| | Vaccination<br>2022<br>Baseline PEP | \$8,250,969 | \$7,078,203 | \$7,148,555 |
| Average cost<br>per human<br>death<br>averted | No vaccination<br>No PEP | Ref | Ref | Ref |
| | No vaccination<br>Baseline PEP | \$1,950 | \$2,018 | \$1,953 |
| | Vaccination<br>2021<br>Baseline PEP | \$1,355 | \$1,651 | \$1,323 |
| | Vaccination<br>2022<br>Baseline PEP | \$1,475 | \$1,510 | \$1,377 |

<sup>†</sup> Total human rabies deaths; we assume only 10% of these would be reported
\* PEP = post-exposure prophylaxis. The baseline PEP assumption is that 80% (outside Ouest Department) or 85% (Ouest Department) of humans exposed to a suspected rabid dog receive PEP, and that 1/5 suspected exposures are true rabies exposures. In people with true rabies exposures, PEP reduces the probability of death. PEP coverage is assumed to remain fixed throughout 2021-2025 in all scenarios.
\*\* Baseline scenario uses input values as in Table S1. High turnover scenario uses a higher-than-baseline values for dog birth rate and mortality (which define dog turnover rate) and lower $R_0$ (number dogs infected per infectious dog). Low turnover scenario uses lower values defining dog turnover but a higher $R_0$ value (Supplementary Appendix Table S2).

## DISCUSSION

Dog rabies vaccination campaigns may be disrupted for a variety of reasons including lack of funding, political instability, or natural disasters; most recently, the COVID-19 pandemic has interrupted dog vaccination in Haiti as well as other countries ^12^. We adapted a mathematical model of dog rabies transmission to assess the urgency of resuming dog vaccination campaigns in Haiti by comparing the human health and cost implications if vaccination is resumed in 2021 compared to 2022. In Ouest Department, home of the capital city and over four million people, the last dog vaccination campaign occurred in 2018. Model estimates and current surveillance data for this region show a 5- to 8-fold increase in dog rabies cases starting in the fall of 2020. In the absence of resumed dog rabies vaccination programs, a dog-mediated human rabies epidemic is predicted to begin in 2021. Implementing a dog vaccination campaign in 2021, compared to delaying until 2022, could save 285 human lives and prevent 6,541 human rabies exposures. Resuming dog vaccination in 2021 compared to 2022 would also require 27,714 fewer courses of PEP, resulting in cost savings of $284,794. Although these benefits were somewhat diminished in sensitivity analyses of different parameters for transmission and dog population turnover, vaccinating in 2021 was still predicted to save at least 107 lives and have similar average cost-effectiveness compared to delaying until 2022.

Dog vaccination is recognized as a cost-effective intervention to prevent human rabies deaths ^1-3^. In this study, we estimated that dog vaccination would be cost saving over a 5-year time frame if 3 or more people receive PEP per true rabies exposure. If PEP is used very efficiently, providing PEP alone might appear more cost-effective than dog vaccination over the short 5-year time frame shown here. However, substantially more people would need to receive PEP for exposures to rabid dogs. This increased demand for PEP would need to be considered in PEP supply forecasts. Furthermore, this study does not consider the long-term effects of eliminating canine rabies, which would greatly reduce the need for PEP.

Previous studies have shown that vaccination campaigns must be repeated annually for elimination to be achieved, particularly when there is heterogeneity in vaccination coverage ^7,21^. Though Haiti made great progress in rabies control from 2013-2019, our results show how fragile such progress can be until elimination is achieved. Although the 2017-2018 vaccination campaign reached approximately 70% coverage throughout the country, the 2019 campaign was not conducted in Ouest Department due to lack of funds. The impacts of this one-year gap would have been minimal if the 2020 vaccination campaign had occurred as planned, but the combination of missed campaigns in 2019 and 2020 now threatens Haiti’s previous gains in rabies control, as shown by both surveillance data and model results.

Multiple researchers are raising concerns about the potential downstream impacts of the COVID-19 pandemic on the control of other diseases including HIV, tuberculosis, malaria, and neglected tropical diseases besides rabies ^22,23^. A previous modeling study predicted that declines in surveillance and paused dog vaccination due to the impacts of COVID-19 could increase dog rabies cases in Peru ^12^. In Haiti, the cancellation of the 2020 dog vaccination campaign was attributable to COVID-19 lockdowns and redirection of funds. However, the impact of the missed 2020 vaccination campaign in Haiti could be mitigated by resuming dog vaccination in 2021, which would save hundreds of lives and thousands of PEP courses compared to even one additional year of delay.

The increase in confirmed dog rabies cases in fall of 2020 in Ouest Department was immediately recognized because of investments in laboratory capacity and real-time electronic rabies surveillance. Since 2018, the Haiti Animal Rabies Surveillance Program has used an app-based case investigation system that allows data to be recorded and shared in real time ^19^. The timing of the increase in confirmed cases detected through surveillance matched that predicted by the model. However, our results suggest that the case detection of rabid dogs is still very low (<1%), particularly in Ouest Department. This may reflect the lower number of rabies investigator staff in Ouest Department (4) compared to outside (33), despite higher predicted numbers of rabid dogs in Ouest Departments for the period assessed.

The model used for this paper contains limitations. First, we assumed that the population within each region (Ouest vs. outside Ouest) has a homogeneous density, despite large variations in population density. We partially corrected for this by removing the lowest-density and most-remote areas from our estimates of population and density. These areas are unlikely to have the dog populations necessary to sustain rabies transmission in the absence of repeated importation ^24,25^. Second, we treated the two regions as independent, and do not consider the movement of dogs and humans between them. This could result in an under-estimation of the human and animal rabies cases that could occur outside of Ouest Department in particular. Many input parameters, such as the human-to-dog ratio and vaccination coverage, are challenging to estimate and may vary substantially within small geographic areas and over time. These parameters were selected based on the best data available, and sensitivity analyses around parameters noted as highly sensitive in previous publications resulted in similar qualitative outcomes.

In summary, we predict that delays in dog vaccination resulting from COVID-19 lockdowns and lack of consistent rabies control funds has brought Haiti to the brink of a potential dog-mediated human rabies epidemic. However, resuming dog rabies vaccination in 2021, compared to delaying until 2022, could save hundreds human lives and reduce the need for PEP over the five years from 2021 to 2025. Despite the immediate threats posed by COVID-19, it is essential that countries continue to invest in control of rabies and other neglected communicable diseases.

## Supporting information

Supplemental data: Excel tool

## Data Availability

All data are available in the manuscript or supplementary materials.

## ACKNOWLEDGMENTS

We would like to acknowledge Benjamin Monroe, Yasmeen Ross, and Natael Fenelon for their contributions to this paper.

## AUTHOR CONTRIBUTIONS

Conceptualization: AK, SJ, MIM, RW

Data curation and parameterization: AK, HCJ, PD, KC, RW

Model design and analysis: AK, SJ, MIM, RW

Wrote first draft of manuscript: AK

Provided edits and feedback on manuscript: SJ, HCJ, PD, KC, MIM, RW

## SUPPORTING INFORMATION

SUPPLEMENTARY APPENDIX: Additional methods and results

SUPPLEMENTARY DATA: Excel modeling tool

