## Supplemental appendix for "The urgency of resuming disrupted dog rabies vaccination campaigns: a modeling and cost-effectiveness analysis"

### Model Parameters and Initialization

Table S1 shows key parameters from the model. Parameters are separated into Ouest Department and the rest of Haiti, as the model was run separately for these two different regions. Additional rabies natural history parameters were the same as for previously published versions of the model (1, 2). The models listed here are those for the baseline scenario.

**Table S1: Model parameters, baseline scenario**

| Variable | Value: Ouest | Value: Non-Ouest | Source | Notes |
|---|---|---|---|---|
| Program area | 1,308 $km^2$ | 4,877 $km^2$ | | Excluding low-density or poorly connected areas; see notes below |
| Human population | 3,794,146 | 4,993,365 | | Excluding low-density or poorly connected areas; see notes below |
| Human population density | 2,900 | 1,024 | | Population/area; excluding low-density or poorly connected areas |
| Human birth rate (per 1,000 pop) | 24 | | (3) | |
| Human life expectancy | 64 years | | (3) | |
| Human-to-Dog ratio at start of burn-in | 20 | 10 | | See notes below |
| Spay/neuter | 0% | | | Assumed |
| PEP coverage | 85% | 80% | (4) | 94% of those with exposure to a confirmed rabid animal and 67% probable started vaccination after counseling |
| Ratio suspected rabies exposures: true rabies exposures (PEP efficiency) | 5 | | (5) | 10:1 is consistent with data from the United States and Haiti (Fig S2), but may not hold in an epidemic situation; values of 1-10x explored in sensitivity analyses |
| Average duration of immunity | 2 years | | (6-8) | Though vaccines used are licensed for 1 year only, this value seems conservative |
| Dog natural life expectancy | 3 years | | (9, 10) | This is if no rabies and no carrying capacity |
| Dog birth rate (per 1,000 dogs) | 650 | | | This corresponds to an observed average life expectancy of ~1.5 years at the end of the burn-in period |
| Rabies $R_0$ dog-to-dog | 1.5 | | (11, 12) | |
| Dog-human transmission rate | $2.34*10^{-5}$ | | (7)<br>(13) | This is the value used in (7); furthermore, the output numbers of human rabies deaths in Haiti are consistent with previous modeled estimates (13). Note this can be conceptualized as (average # humans bitten per rabid dog)/(human population density) |
| Community bite surveillance costs per case investigated | $10.14 (USD) | | (2) | |
| Cost per dog vaccinated | $2.92 (USD) | | | Based on an evaluation of a vaccination campaign in Artibonite Department in 2017 |
| Cost of post-exposure prophylaxis, per person | $72.91 (USD) | | (2) | |

**APPENDIX – SUPPLEMENTAL METHODS AND RESULTS**

**The urgency of resuming disrupted dog rabies vaccination campaigns: a modeling and cost-effectiveness analysis**

Modified values of the dog birth rate, the dog natural life expectancy, and the $R_0$ for dog-dog transmission used in sensitivity analyses are shown in Table S2.

**Table S2: Parameter values modified in sensitivity analyses.**

| | Baseline scenario | "High turnover" scenario | "Low turnover" scenario |
|---|---|---|---|
| Dog birth rate (per 1,000 dogs) | 650 | 800 | 500 |
| Dog natural life expectancy, years | 3.0 | 2.5 | 3.5 |
| $R_0$ dog to dog* | 1.5 | 1.3 | 1.7 |

* $R_0$ is the average number of dogs infected with rabies per infectious (rabid) dog, in a situation where all other dogs are susceptible to infection.

In estimating human and dog population density, we aimed to include only those areas with density and connectivity high enough to maintain rabies transmission in the absence of isolated importation events (14, 15). To define low-density areas, we used a shapefile from the Gates Foundation containing polygons for inhabited areas (16). Using QGIS, we classified each polygon according to the department in which it was located (for polygons crossing department borders, we took the one with greatest overlap). We used the surface area of inhabited areas inside and outside of Ouest Department as our input areas.

We then calculated the population for each polygon based on population raster grids from (17) using the "zonal statistics" command. We used the calculated population of the inhabited areas inside and outside of Ouest Department as our input population values. We assumed a human-to-dog ratio of 20:1 (Ouest Department) and 10:1 (outside Ouest Department) in the absence of rabies. These values are consistent with estimates from other urban areas (14, 18), and were chosen to reflect the greater human density in Ouest Department. Surveys taken in 2014-2015 in 5 departments of Haiti showed human-to-dog ratios of 3-147 humans/dog. Excluding the outlier of 147, the mean estimates were 14.6 in Ouest and 9.7 outside Ouest. Surveys in 16 communities of Croix-des-Bouquets Commune, Ouest Department in 2016 ranged from 4-29 humans per dog.

The model was initialized by adding a small number of cases and running forward until a steady state is reached. In contrast to previous implementations of RabiesEcon, we assumed 5% annual vaccination coverage during this burn-in period, until 2005.

### APPENDIX – SUPPLEMENTAL METHODS AND RESULTS

#### The urgency of resuming disrupted dog rabies vaccination campaigns: a modeling and cost-effectiveness analysis

Model Fit 2005-2020

The model-predicted number of reported cases and the number of rabies exposures for Haiti from 2005-2020 are roughly consistent with observed surveillance data. Though the trends do not fully match, surveillance efforts have been variable over time, and the results are consistently within an order of magnitude of the expected values. Table S3 shows the trends in human rabies deaths detected by surveillance compared to model estimates, as well as model estimates of human rabies exposures, under the baseline parameter set for the years 2005-2020.

**Table S3: Model predicted reported rabies deaths, total rabies deaths, and total rabies exposures for 2005-2020, compared to surveillance data on rabies deaths per year.**

| Year[1] | Assumed Dog Vaccination Coverage | Surveillance-Detected deaths to rabies | Model estimate: reported[2] human rabies deaths | Model estimate: total human rabies deaths | Model estimate: total human rabies exposures |
|---|---|---|---|---|---|
| 2005 | 25% | - | 12 | 124 | 2439 |
| 2006 | 25% | - | 6 | 62 | 1153 |
| 2007 | 30% | - | 3 | 34 | 656 |
| 2008 | 30% | - | 2 | 23 | 467 |
| 2009 | 35% | - | 2 | 21 | 439 |
| 2010 | 0% | 1 | 3 | 28 | 722 |
| 2011 | 0% | 13 | 17 | 169 | 4619 |
| 2012 | 35% | 2 | 45 | 447 | 8901 |
| 2013 | 0% | - | 13 | 132 | 2331 |
| 2014 | 0% | - | 11 | 105 | 2316 |
| **After the implementation of Haiti Animal Rabies Surveillance Program (HARSP) and PAHO Human Rabies Vaccine Training Program** | | | | | |
| 2015 | 50% | 5 | 13 | 127 | 2402 |
| 2016 | 10% | 7 | 3 | 30 | 558 |
| 2017 | 0% (Ouest)<br>70% (Non-Ouest) | 9 | 4 | 37 | 881 |
| 2018 | 70% (Ouest)<br>0% (Non-Ouest) | 4 | 6 | 56 | 1185 |
| 2019 | 0% (Ouest)<br>70% (Non-Ouest) | 2 | 1 | 9 | 166 |
| 2020 | 0% | 1 | 2 | 15 | 453 |
| **Overall (2005-2020)** | | **44** | **142** | **1,419** | **29,688** |
| Pre-HARSP (2005-2014) | | 16 | 114 | 1,145 | 24,043 |
| Post-HARSP (2015-2020) | | 28 | 27 | 274 | 5,645 |

*[1]Vaccination was assumed to begin halfway through the year. [2]Assuming 10% case detection (see Methods).*

The three parameter sets used for sensitivity analyses showed similar trends over time through 2020, as can be shown in Figures S1-S3.

**APPENDIX – SUPPLEMENTAL METHODS AND RESULTS**

**The urgency of resuming disrupted dog rabies vaccination campaigns: a modeling and cost-effectiveness analysis**

Figure S1 shows the number of human rabies deaths per year from 2005-2020 estimated by the model for both Ouest Department and outside Ouest Department. These estimates are roughly consistent with the estimate of 130 human rabies deaths per year in Haiti (13).

**Fig S1: Model estimates of human rabies deaths per year, 2005-2020**

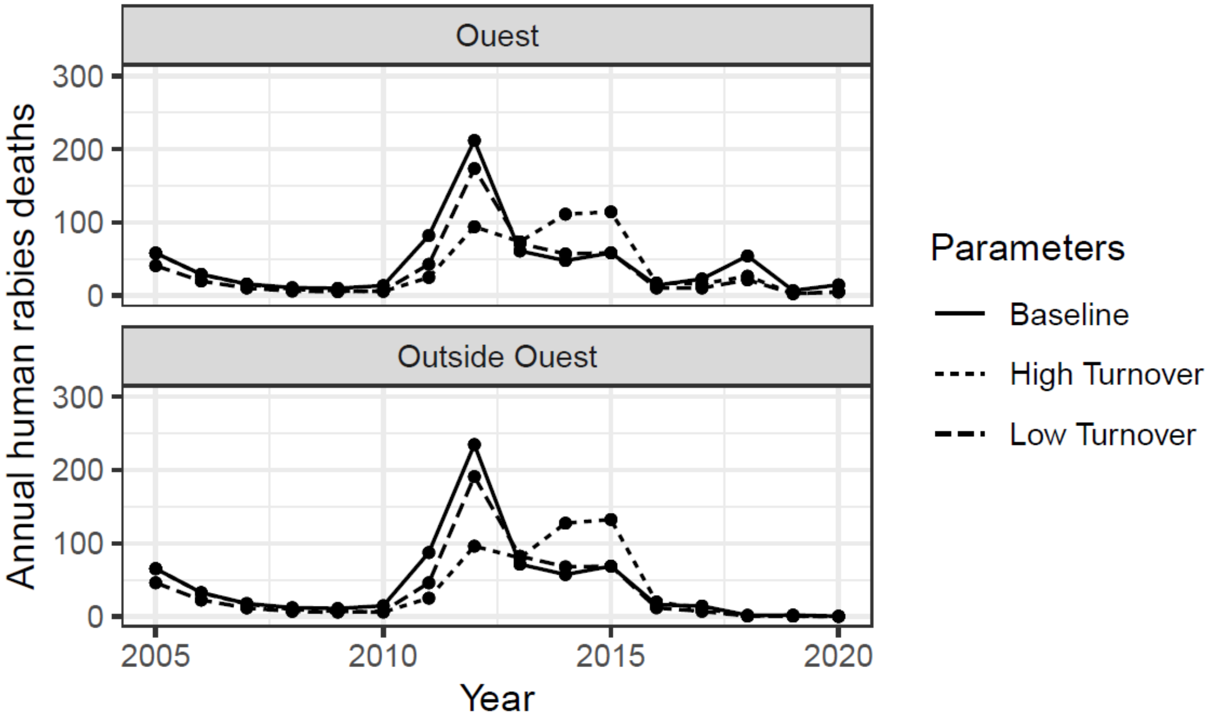


Figure S2 shows the number of human rabies exposures estimated by the model per year from 2005-2020 for both Ouest Department and outside of Ouest Department. Data from Haiti showed 1,111 total individuals initiated PEP during the first 17 weeks of 2016, including individuals with exposures to animals of unknown rabies status. Assuming 1111*52/17=3398 individuals initiated PEP during all of 2016, and comparing with the baseline modeled estimate of 558 true human rabies exposures, suggests a ratio of 6.1 PEP courses per true rabies exposure. Data from the first 48 weeks of 2020 showed 3566 initiated PEP. Assuming 3566*52/48=3863 individuals initiated PEP during all of 2020, and comparing with the baseline modeled estimates of 453 true human rabies exposures, suggests a ratio of 8.5 PEP courses per true rabies exposure.

**The urgency of resuming disrupted dog rabies vaccination campaigns: a modeling and cost-effectiveness analysis**

**Fig S2: Model estimates of human rabies exposures per year, 2005-2020**

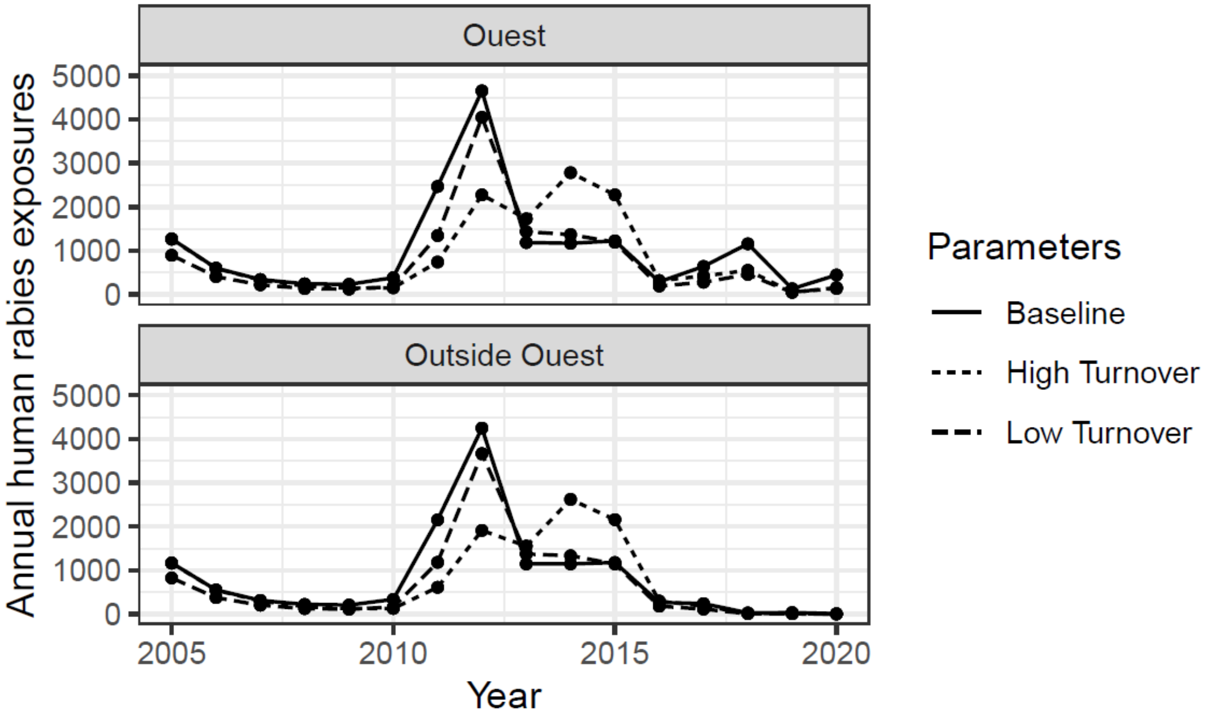


Figure S3 shows the number of canine rabies cases per year from 2005-2020 for both Ouest Department and outside of Ouest Department, divided by the size of the canine population at the end of the given year. This is not exactly incidence, considering that population size is measured at the end of the year, and the high turnover of the dog population throughout the year. Nevertheless, the estimates appear similar to data from other settings that generally report below 0.5-2% of dogs developing rabies per month, as summarized in (19). One previous study reported 10% of dogs found dead in Haiti were positive for rabies (20). This further validates that our model estimated total human rabies deaths and human exposure are within a reasonable range compared to the observed surveillance data and previously published studies.

**The urgency of resuming disrupted dog rabies vaccination campaigns: a modeling and cost-effectiveness analysis**

**Fig S3: Model estimates of annual canine rabies cases divided by canine rabies population at year end, 2005-2020**

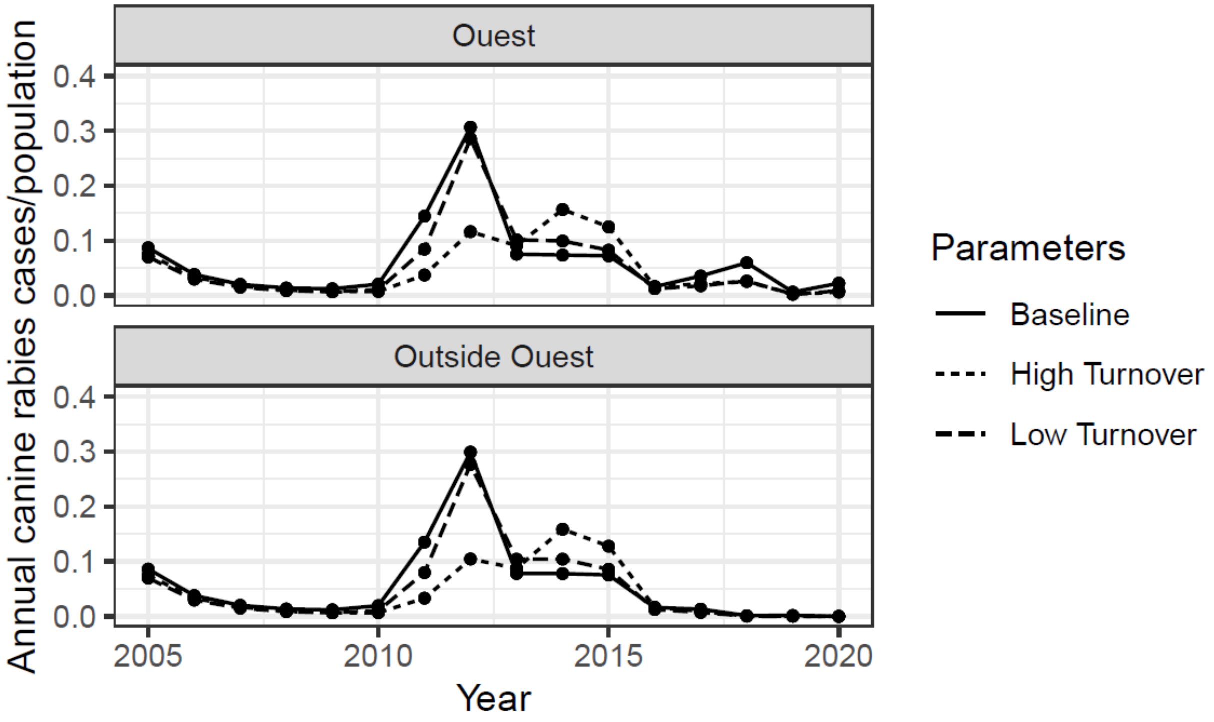

APPENDIX – SUPPLEMENTAL METHODS AND RESULTS

**The urgency of resuming disrupted dog rabies vaccination campaigns: a modeling and cost-effectiveness analysis**

Supplemental Results

Table S4 compares animal surveillance data with model estimates of rabid dogs. The estimated case detection is low: 0.1-0.6% in Ouest Department, compared to 0-3.9% outside Ouest Department. Investigations focus primarily on dog bites. The higher case detection rates outside Ouest might be explained in part by the higher number of rabies investigators outside Ouest Department (4 investigators in Ouest Department vs 33 outside, despite higher model estimates of rabid dogs in Ouest Department than outside for the time period depicted here).

**Table S4: Surveillance data on suspected and confirmed rabid animals, 2018-2020**

| Time period | Investigations | Tested* | Confirmed | Percentage of investigations confirmed positive | Model estimated rabid dogs | Case detection rate (confirmed/model estimate) |
|---|---|---|---|---|---|---|
| **Ouest Department** | | | | | | |
| 2018 Fall | 382 | 7 | 3 | 0.8% | 2,003 | 0.1% |
| 2019 Spring | 401 | 5 | 3 | 0.7% | 577 | 0.5% |
| 2019 Fall | 318 | 2 | 1 | 0.3% | 604 | 0.2% |
| 2020 Spring | 292 | 3 | 1 | 0.3% | 1,154 | 0.1% |
| 2020 Fall[1] | 284 | 11 | 8 | 2.8% | 3,030 | 0.3% |
| **Outside Ouest Department** | | | | | | |
| 2018 Fall | 946 | 13 | 7 | 0.7% | 355 | 2.0% |
| 2019 Spring | 1,177 | 29 | 17 | 2.5% | 667 | 2.6% |
| 2019 Fall | 886 | 8 | 6 | 0.7% | 193 | 3.1% |
| 2020 Spring | 589 | 7 | 0 | 0% | 77 | 0% |
| 2020 Fall[1] | 398 | 3 | 1 | 0.3% | 114 | 0.9% |

*Only investigated animals with high suspicion of rabies are tested.

[1]Data are included through November 19, 2020 only, but model output includes the entire 6-month period.

**APPENDIX – SUPPLEMENTAL METHODS AND RESULTS**

**The urgency of resuming disrupted dog rabies vaccination campaigns: a modeling and cost-effectiveness analysis**

**Figure S4: Historical and projected canine rabies cases in Haiti, 2015-2025, and potential impact of delaying restarting dog rabies vaccination until either 2021 or 2022.**

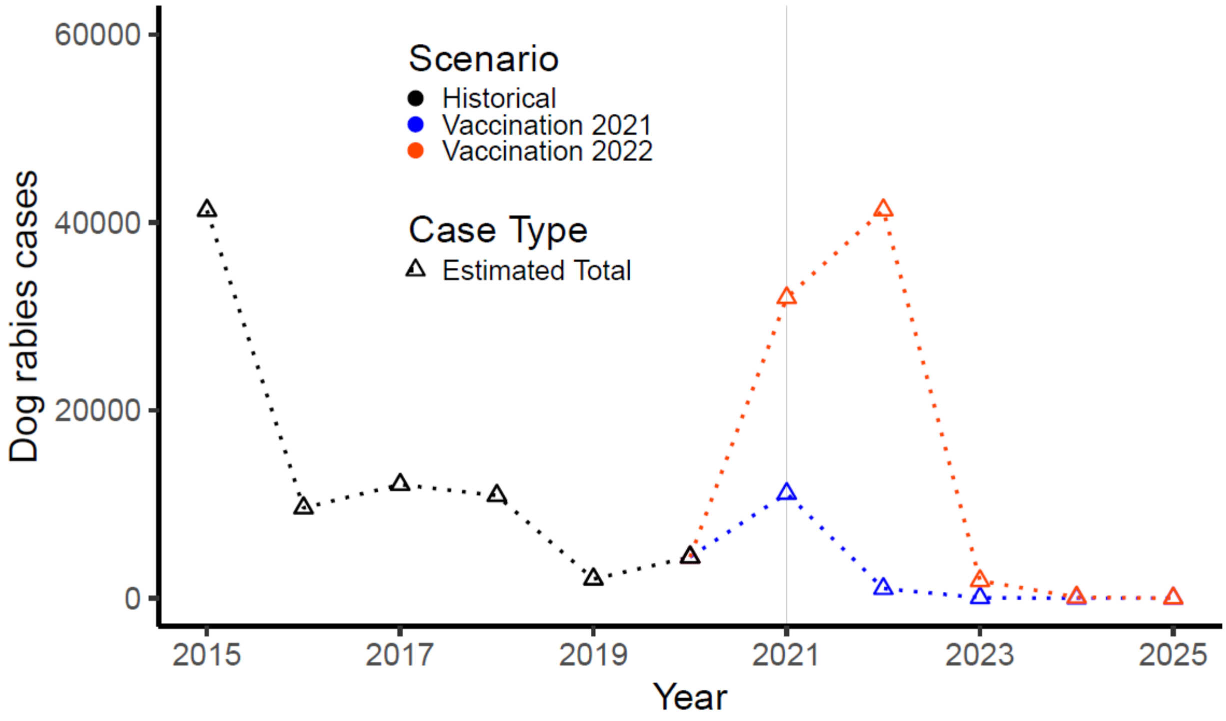


* Triangles (connected by dotted lines) represent model estimates of total dog rabies cases. Colors compare historical data and estimates for 2015-2020 (black) with model predictions if vaccination campaigns resume in 2021 (blue) or 2022 (orange). See Figure 2 in the main text for similar estimates of human rabies deaths.

### APPENDIX – SUPPLEMENTAL METHODS AND RESULTS

### The urgency of resuming disrupted dog rabies vaccination campaigns: a modeling and cost-effectiveness analysis
